# SPECIAL CARE BABY UNIT NEONATAL DISEASE OUTCOMES IN A TERTIARY HOSPITAL IN NIGERIA: 2-YR RETROSPECTIVE CROSS-SECTIONAL ANALYSIS

**DOI:** 10.1101/2024.10.02.24314811

**Authors:** Udochukwu Godswill Anosike, Ugochukwu Godson Amalahu, Chijioke Amara Ezenyeaku, Chika Florence Ubajaka, Anokwulu Ifeanyi Osmond, Chiamaka Sandra Nsude, Joseph Moses Adeniyi, Chinemerem Okonkwo, Uzoma Love Nwajinka, Malachy Echezona DivineFavour, Chukwuemelie Darlington Okeke, Chidozie Valentine Akwiwu-Uzoma

## Abstract

**BACKGROUND:** Neonatal diseases contribute significantly to global under-five mortality. The highest neonatal mortality rate in sub-Saharan Africa can be traced to Nigeria. This study aims to evaluate the outcomes of neonatal admissions in a select tertiary hospital in Nigeria.

**METHODS:** A retrospective analysis of data collected on 656 neonates admitted in the special care baby unit of Nnamdi Azikiwe University Teaching Hospital (NAUTH), Nigeria over a period of 2 years (January 2021and December 2022). Descriptive analysis and inferential statistics were done at p<0.05 using SPSS version 25.

**RESULTS:** Median age at presentation was 4 hours (interquartile range 0.5,24) hours. The Median duration of hospital stay was 6 days (interquartile range 3,11). The commonest morbidities were perinatal asphyxia (n=295/656; 45.0%) and prematurity (n=158/656; 24%); while congenital anomalies (n=22/47; 46.8%), perinatal asphyxia (n=73/295; 24.7%) and prematurity (n=35/158; 22.2%) had the highest case fatality rates. Gestational age at birth, duration of hospital stay, place of delivery, and mode of delivery were the variables determined to be statistically associated with outcome of care.

**CONCLUSION:** This study showed a mortality of 22.9% (n=150/656) in our study area with perinatal asphyxia (48.7%; n=73/150), prematurity (24.0%; n=36/150), congenital anomalies (11.3%; n=22/150), and neonatal sepsis (6.7%; n=10/150) as the primary causes. This work highlights the need for emergency care of critically ill newborns through financing transition from special care baby unit to neonatal intensive care unit across tertiary institutions in Nigeria.

**KEY MESSAGES:** *What is already known on this topic:*

- Poor outcome of neonatal disease is associated with lack of quality care at birth or skilled care and treatment immediately after birth and in the first days of life.
- Premature birth, birth asphyxia, neonatal infections and congenital anomalies are the prominent causes of neonatal deaths.

*What this study adds:*

- Case fatality rate, mortality rate and duration of hospital stay were the primary outcomes assessed in a spectrum of neonatal disease.
- Higher mortality was recorded among neonates within the first 7days of hospital stay.

*How this study might affect research, practice, or policy (implications):*

- Our work informs the need for transition from special care baby unit to neonatal intensive care unit across tertiary institutions in Nigeria to enable comprehensive care for critically ill neonates or neonates with severe neonatal morbidities.
- Creates awareness on the need for incorporation of processes that enable safeguarding of neonatal hospital records for tracking of trends and research.

## 1. INTRODUCTION

Neonatal diseases are a significant global health issue, particularly in high-risk populations, such as preterm babies, and low birth weight neonates. The leading causes of neonatal morbidity and mortality include prematurity and low birth weight, perinatal asphyxia, sepsis and infection.^1,2^ Most of these deaths (almost 99%) occur in developing countries (low and middle-income countries).^3^ In sub-Saharan Africa, neonatal diseases contribute to 39% of under-five mortality.^4^ Nigeria was ranked by World Health Organization as the country with the second highest number of newborn deaths in 2020, averaging about 271,000 deaths.^5^ According to Statista 2019, neonatal disorders (25.27%) were the major cause of under-five mortality in Nigeria.^6^ Between 2016 and 2017, the neonatal mortality rate (NMR) in Nigeria was 37 per 1000 live births.^7^ In other sub-Saharan African countries, NMR, as reported, are Kenya (2015) - 22/1000 live births ^8^, Ethiopia (2016) - >35/1000 live births ^9^, and Cameroon (2017) - 21/1000 live births.^10^ The magnitude of neonatal mortality across tertiary hospitals in various regions and states in Nigeria are as follows – 12.2% in rivers state^11^, and 10.5% in North Central Nigeria (45/428)^12^. This study therefore provides extensive insights into the distribution of neonatal diseases, and the outcomes of admissions in a selected Nigerian hospital.

## 2. METHODS

### 2.1. STUDY AREA

This study was conducted in the special care baby unit of Nnamdi Azikiwe University Teaching Hospital, (NAUTH) Nnewi. Nnewi is a commercial and industrial city in Anambra State, Southeastern Nigeria. The special care baby unit of Nnamdi Azikiwe University Teaching Hospital, Nnewi was established in May 1998.^13^ It consists of 3 wards (for in-born babies, out-born/referred babies, and isolation), and contains 17 cots, 1 infant radiant warmer, 5 incubators, 2 oxygen cylinders, 5 phototherapy units, 1 apnea monitor and 4 resuscitation kits.

### 2.2. STUDY DESIGN

This study was a retrospective descriptive cross-sectional study.

### 2.3. STUDY POPULATION

The study population was newborn babies admitted to the special care baby unit, NAUTH, Nnewi.

#### 2.3.1. INCLUSION CRITERIA

All neonates between 0 and 28 days who presented between January 2021 and December 2022.

#### 2.3.2. EXCLUSION CRITERIA

Neonates with incomplete data on health records, infant of mothers with sickle cell disease, and stable term neonates admitted to special care baby unit.

### 2.4. SAMPLE SIZE DETERMINATION

This study was a whole population study. The minimum sample size (n) for this study was calculated using Cochran’s sample size (n) formula for a known finite population (N).

Where n is Cochran’s sample size, given as:

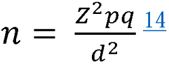

Where, n = minimum sample size.

d= 0.05, desired level of precision (i.e., 5% margin of error)
p = 0.21, using 21% (prevalence of perinatal asphyxia in a tertiary institution) ^15^
q = 1 – p, which is 0.79
z = 1.96, standard normal deviate at 95% confidence interval (CI)

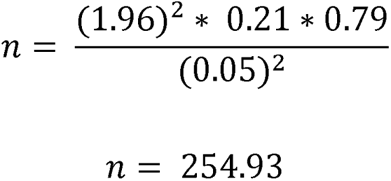

The minimum sample size, as calculated for the two years studied was approximately 255.

### 2.5. SAMPLING TECHNIQUE

All folders of neonates that met the inclusion criteria within the study period were retrieved for data collection.

### 2.6. STUDY INSTRUMENT

A checklist (proforma) was used to obtain information on the patient’s profile which included: the mother’s age, marital status, occupation, level of education, parity, sex, birth weight, gestational age at birth, age at presentation, diagnosis at presentation, duration of hospital stay, and outcome.

### 2.7. TRAINING OF RESEARCH ASSISTANTS

Ten (10) medical students in their 5^th^ year of study were recruited and trained for a day by the principal researchers as research assistants. They were trained on how to extract the required information from folders using the checklist. The success of the training was assessed with the preliminary data collected. To further ensure data quality, the principal researcher monitored the data collection. There were meetings of the data collection team at the end of each collection day to collate the data.

### 2.8. DURATION OF STUDY

The study lasted for 3 months, starting from May to July 2023.

### 2.9. DATA COLLECTION METHODS

With permission from the medical records department and Special Care Baby Unit NAUTH, the medical records of the neonates were identified, retrieved, studied, and the required data were extracted from all that met the inclusion criteria using the checklist.

### 2.10. DATA MANAGEMENT

#### 2.10.1. MEASUREMENT OF VARIABLES

The main outcome variables for this study were – fatality rate of cases, neonatal mortality rate, duration of hospital stay.

Other variables that affected the outcomes include birth weight, gestational age at birth, place of and delivery.

#### 2.10.2. STATISTICAL DATA ANALYSIS

Data cleaning was done using Microsoft office Excel, version 2021 (Microsoft Corporation, Redmond, WA, USA). Duplicated and incomplete data were removed. Further analysis of data was done using Statistical Package for Social Sciences (SPSS) version 25 (IBM Corp., Armonk, NY, USA). All continuous variables were tested for normality using Shapiro-Wilks’s test and found not to be normally distributed. Thus, median, and interquartile ranges were used as measures of central tendency for them. Quantitative variables were further divided into categories. Categorical variables were summarized using tables of frequencies and percentages. The Fisher’s exact test with Bonferroni correction was used for testing the significance of associations between outcome of hospital admissions and various variables. The level of significance was set at P<0.05.

### 2.11. ETHICAL CONSIDERATIONS

Approval was obtained from the Nnamdi Azikiwe University Teaching Hospital Health Research Ethics Committee (NAUTHHREC) through the Head of the Department of Community Medicine, Nnamdi Azikiwe University. Permission was also obtained from the Pediatrics Head of Department, Special Care Baby Unit and Head of the Department of Medical Records. The data extracted from the case notes was treated with utmost confidentiality, and patient names and other details that could lead to their identification were excluded to maintain anonymity.

## 3. RESULTS

The total number of neonatal admissions between 2021 and 2022 was 973. 32% of health records (n=312/973) were not assessed due to the missing health records and incompleteness of data as required by proforma. 5 out of 661 assessed health records were excluded – stable term neonates (4) and infant of mother with sickle cell disease (1).

**Table 1.1:**
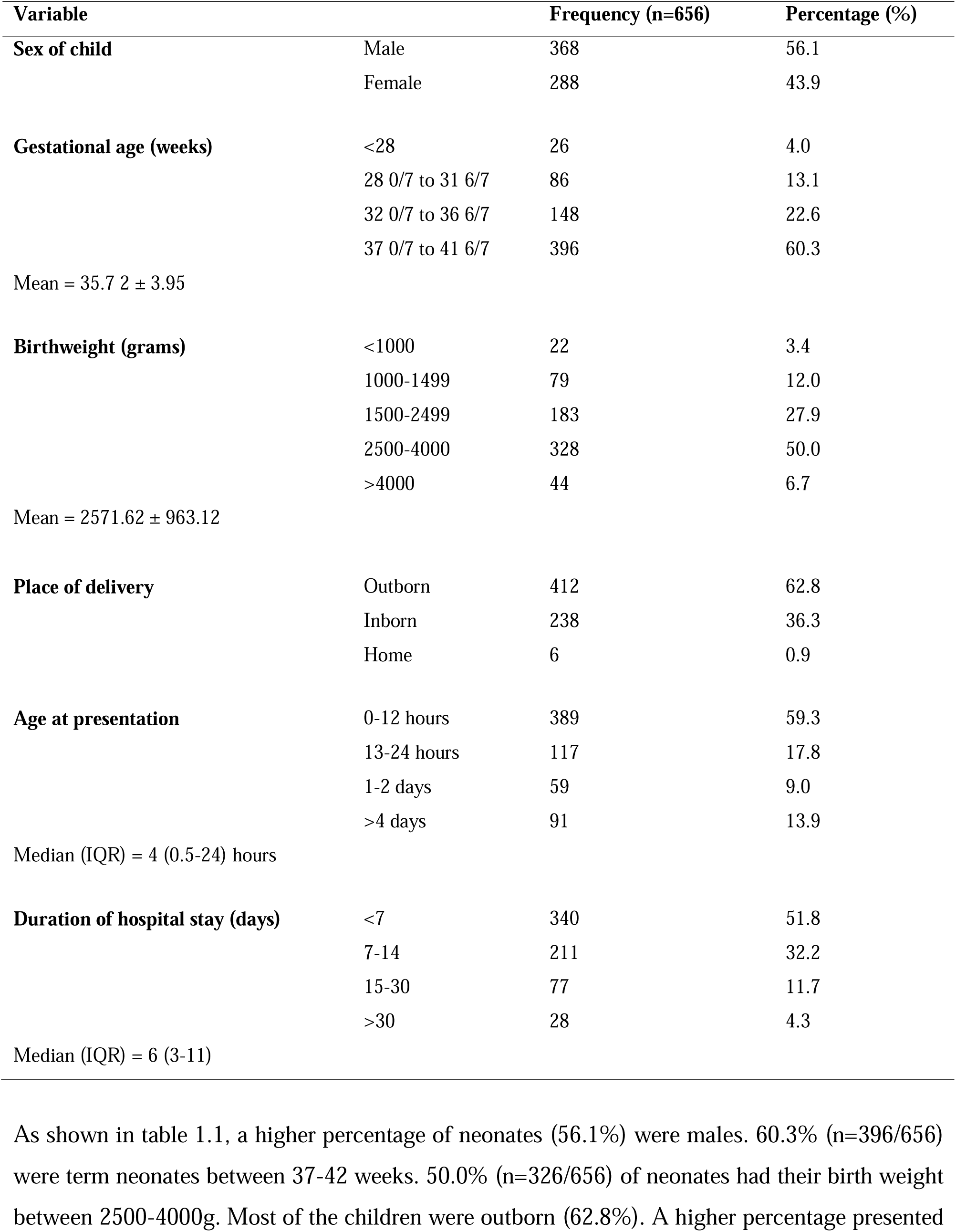
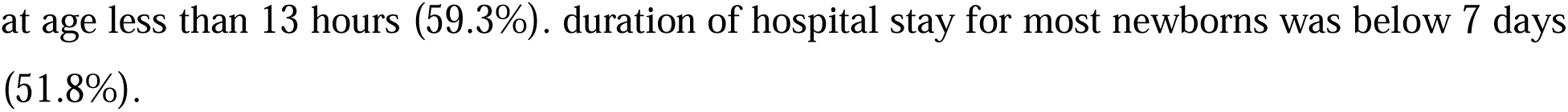
Characteristics of neonates admitted to special care baby unit NAUTH.

**Table 1.2:** Maternal characteristics.

| Variable |  | Frequency (n=656) | Percentage (%) |
| --- | --- | --- | --- |
| **Age of mother (years)** | 18-29 | 340 | 51.8 |
|  | 30-39 | 271 | 41.3 |
|  | >39 | 45 | 6.9 |
|  | Median (IR) = 29 (25-34) years |  |  |
| **Marital status** | Single | 23 | 3.5 |
|  | Married | 633 | 96.5 |
| **Parity** | Primipara | 223 | 34.0 |
|  | Multipara | 433 | 66.0 |
| **Educational level** | Tertiary | 212 | 32.3 |
|  | Secondary | 388 | 59.1 |
|  | Primary | 37 | 5.6 |
|  | No formal | 19 | 2.9 |
| **Occupation** | Unemployed | 50 | 7.6 |
|  | Trader | 366 | 55.8 |
|  | Student | 45 | 6.9 |
|  | Professional | 42 | 6.4 |
|  | Housewife | 91 | 13.9 |
|  | Civil servant | 56 | 8.5 |
|  | Artisan | 6 | 0.9 |
| **Religion** | Muslim | 3 | 0.5 |
|  | Christian | 653 | 99.5 |
| **Ethnicity** | Igbo | 653 | 99.5 |
|  | Hausa | 3 | 0.5 |
| **Mode of delivery** | Spontaneous vaginal delivery | 388 | 59.1 |
|  | Emergency caesarean section | 204 | 31.1 |
|  | Elective caesarean section | 60 | 9.1 |
|  | Assisted vaginal delivery | 4 | 0.6 |
| **Premature rupture of membrane** | Yes | 114 | 17.4 |
|  | No | 542 | 82.6 |
| **Peripartum fever** | Yes | 162 | 24.7 |
|  | No | 494 | 75.3 |
| **Dexamethasone administration** | Yes | 143 | 21.8 |
|  | No | 513 | 78.2 |

**Table 1.3:** Distribution of neonatal diseases in special care baby unit NAUTH.

| Diagnosis at presentation | Frequency (n-656) | Percentage (%) |
| --- | --- | --- |
| Congenital anomalies | 47 | 7.2 |
| Macrosomia | 26 | 4 |
| Neonatal jaundice | 34 | 5 |
| Neonatal sepsis | 63 | 9.6 |
| Perinatal asphyxia | 295 | 45 |
| Prematurity | 158 | 24 |
| Respiratory distress syndrome | 17 | 2.6 |
| Transient tachypnea of the newborn | 7 | 1.1 |
| Vitamin K deficiency bleeding | 9 | 1.4 |

**Table 1.4:** Outcome of admission for neonatal diseases in the special care baby unit NAUTH.

| Outcome of care | Frequency (n=656) | Percentage (%) |
| --- | --- | --- |
| DAMA | 32 | 4.9 |
| Died | 150 | 22.9 |
| Discharged | 454 | 69.2 |
| Transferred | 20 | 3.0 |

DAMA = Discharged against medical advice

**Table 1.5:** Relationship between diagnosis at presentation and outcome of care (n=656)

| Diagnosis at presentation | Outcome n (%) |  |  |  | F(p) |
| --- | --- | --- | --- | --- | --- |
|  | Transferred | Discharged | Died | DAMA |  |
| Congenital anomalies | 7(14.9) | 17(36.2) | 22(46.8) | 1(2.1) | 89.750 (<0.001) |
| Macrosomia | 0 | 23(88.5) | 2(7.7) | 1(3.8) |  |
| Neonatal jaundice | 0 | 22(64.7) | 6(17.6) | 6(17.6) |  |
| Neonatal sepsis | 4(6.3) | 47(74.6) | 10(15.9) | 2(3.2) |  |
| Perinatal asphyxia | 6(2.0) | 205(69.5) | 73(24.7) | 11(3.7) |  |
| Prematurity | 0 | 114(72.1) | 35(22.2) | 9(5.7) |  |
| Respiratory distress syndrome | 2(11.8) | 11(64.7) | 2(11.8) | 2(11.8) |  |
| Transient tachypnea of the newborn | 1(14.3) | 6(85.7) | 0 | 0 |  |
| Vitamin K deficiency bleeding | 0 | 9(100) | 0 | 0 |  |

**Table 1.6:** Analysis of maternal factors influencing burden of neonatal diseases in special care baby unit NAUTH (n=656)

| Variables |  | Outcome n (%) |  |  |  | F(p) |
| --- | --- | --- | --- | --- | --- | --- |
|  |  | Transferred | Discharged | Died | DAMA |  |
| Age of mother (years) | 18-29 | 10(2.9) | 234(68.8) | 74(21.8) | 22(6.5) | 5.16(0.49) |
|  | 30-39 | 8(3) | 191(70.5) | 63(23.2) | 9(3.3) |  |
|  | >39 | 2(4.4) | 29(64.4) | 13(28.9) | 1(2.2) |  |
| Marital status | Single | 1(4.3) | 13(56.5) | 8(34.8) | 1(4.3) | 2.78(0.37) |
|  | Married | 19(3) | 441(69.7) | 142(22.4) | 31(4.9) |  |
| Level of education | Tertiary | 8(3.8) | 145(68.4) | 47(22.2) | 12(5.7) | 4.80(0.81) |
|  | Secondary | 10(2.6) | 271(69.8) | 88(22.7) | 19(4.9) |  |
|  | Primary | 2(5.4) | 24(64.9) | 11(29.7) | 0 |  |
|  | No formal | 0 | 14(73.7) | 4(21.1) | 1(5.3) |  |
| Occupation | Unemployed | 3(6) | 33(66) | 13(26) | 1(2) | 5.69(0.15) |
|  | Trader | 13(3.6) | 257(70.2) | 77(21) | 19(5.2) |  |
|  | Student | 2(4.4) | 30(66.7) | 10(22.2) | 3(6.7) |  |
|  | Professional | 1(2.4) | 27(64.3) | 12(28.6) | 2(4.8) |  |
|  | Housewife | 0 | 63(69.2) | 23(25.3) | 5(5.5) |  |
|  | Civil servant | 1(1.8) | 42(75) | 11(19.6) | 2(3.6) |  |
|  | Artisan | 0 | 2(33.3) | 4(66.7) | 0 |  |
| Parity | Primipara | 7(3.1) | 150(67.3) | 52(23.3) | 14(6.3) | 1.68(0.64) |
|  | Multipara | 13(3) | 304(70.2) | 98(22.6) | 18(4.2) |  |
| Mode of delivery | Spontaneous vaginal delivery | 12(3.1) | 259(66.8) | 97(25) | 20(5.2) | 22.37(0.01) |
|  | Emergency caesarean section | 7(3.4) | 144(70.6) | 43(21.1) | 10(4.9) |  |
|  | Elective caesarean section | 1(1.7) | 51(85) | 8(13.3) | 0 |  |
|  | Assisted vaginal delivery | 0 | 0 | 2(50) | 2(50) |  |
| Premature rupture of membrane | Yes | 1(0.9) | 78(68.4) | 30(26.3) | 5(4.4) | 2.69(0.44) |
|  | No | 19(3.5) | 376(69.4) | 120(22.1) | 27(5) |  |
| Peripartum fever | Yes | 7(4.3) | 118(72.8) | 30(18.5) | 7(4.3) | 3.52(0.31) |
|  | No | 13(2.6) | 336(68) | 120(24.3) | 25(5.1) |  |
| **Dexamethasone administration** | Yes | 1(0.7) | 89(62.2) | 46(32.2) | 7(4.9) | 11.19(0.01) |
|  | No | 19(3.7) | 365(71.2) | 104(20.3) | 25(4.9) |  |

**Table 1.7:**
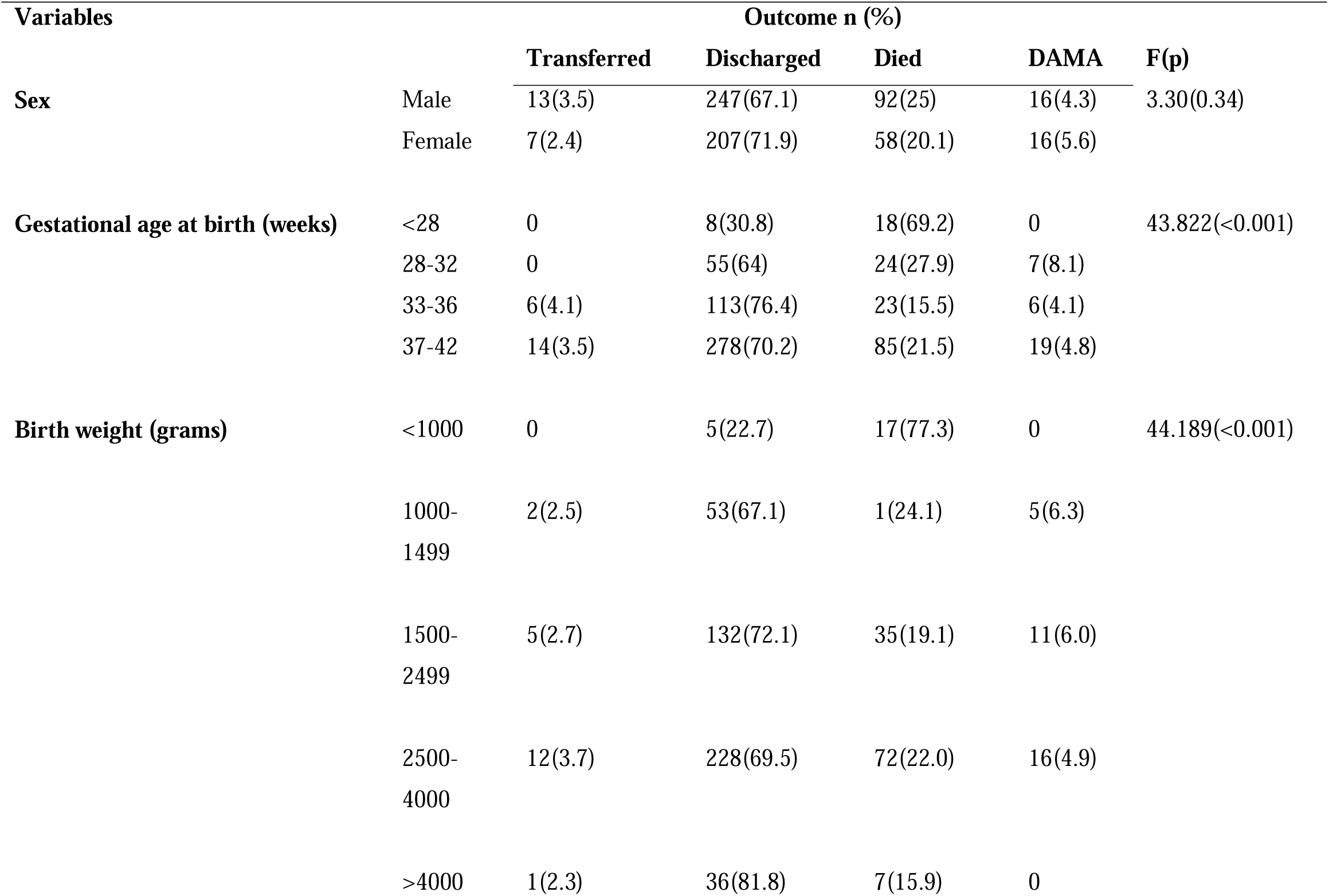
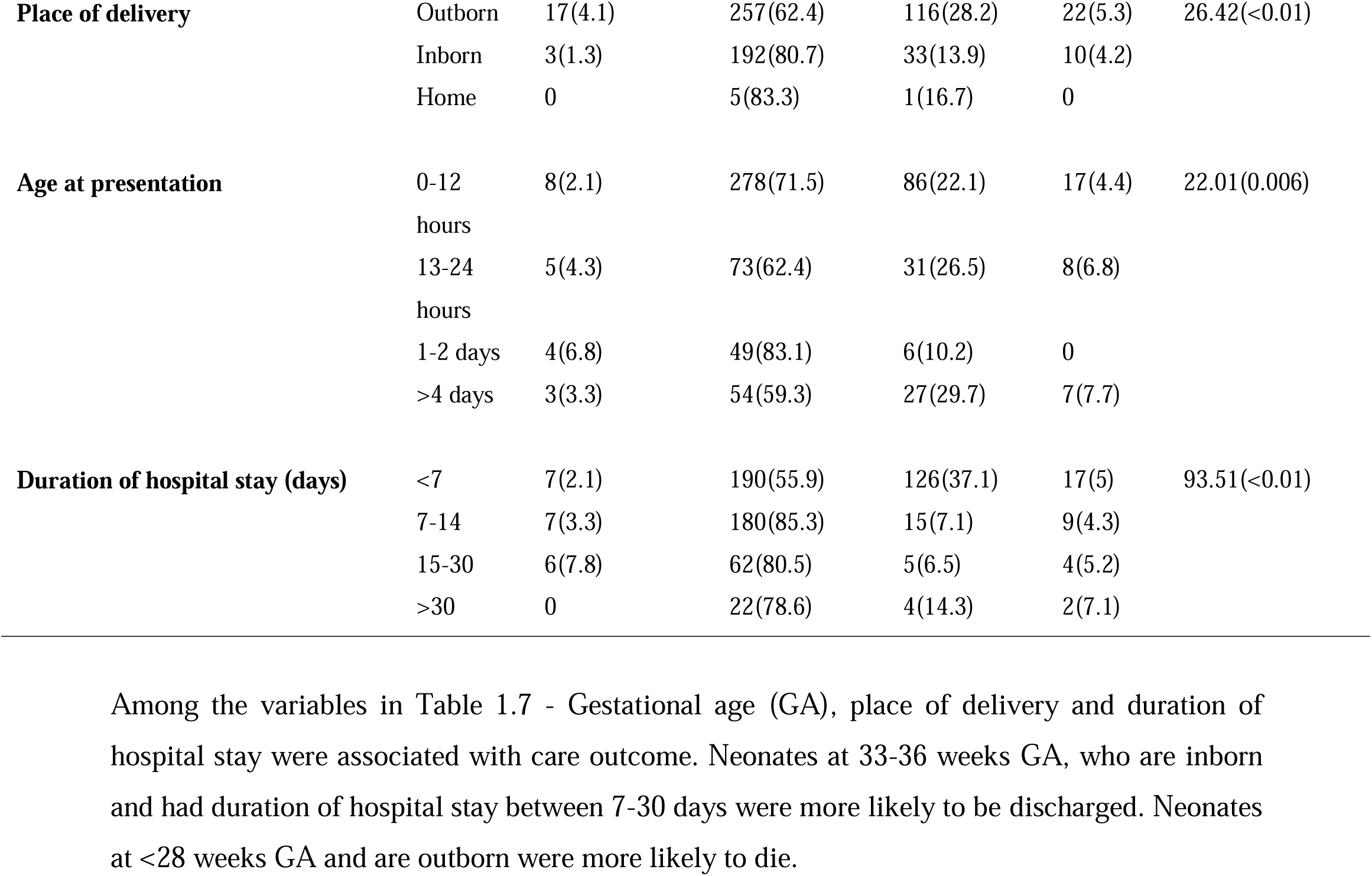
Analysis of neonatal factors influencing outcomes in special care baby unit NAUTH (n=656)

## 4. DISCUSSION

Findings from this study showed a high burden of neonatal morbidity and mortality in hospitalized newborns (656) admitted to special care baby unit in NAUTH, Anambra State with a mortality of 22.9% (n=150/656). This contrasts with a similar study carried out in our study area by Ezechukwu et al which reported a mortality of 19.4% (n=166/854).^13^ This demonstrates a 15.2% increase in neonatal mortality since 2001. Reports from other studies showed lower mortality in tertiary institutions in rivers state (12.2%) ^11^, and 10.5% in North Central Nigeria ^12^

The primary causes of mortality were perinatal asphyxia (48.7%; n=73/150), prematurity (21.3%; n=35/150), congenital anomalies (11.3%; n=22/150), and neonatal sepsis (6.7%; n=10/150). These findings align with a study conducted at ESUTH, which also identified perinatal asphyxia as the predominant cause of mortality.^16^ The highest case fatality rate in this study was recorded amongst neonates with congenital anomalies (n=22/47; 46.8%), perinatal asphyxia (n=73/295; 24.7%) and prematurity (n=35/158; 22.2%). In contrast, other studies reported neonates with respiratory distress syndrome ^17^, neonatal sepsis ^11^ as morbidities with the highest case fatality rate.

Prematurity, perinatal asphyxia, neonatal sepsis, neonatal jaundice, congenital anomalies as reported by Ezechukwu et al 2005^13^ have remained the prevalent disorders in NAUTH. Our study indicate that perinatal asphyxia was the leading cause of neonatal admissions, accounting for 45.0% of cases. Additionally, prematurity, neonatal sepsis, congenital anomalies, and neonatal jaundice contributed to 24.8%, 9.6%, 7.2% and 5.2% of admissions, respectively. These results are consistent with findings from other medical centers across Nigeria.^16, 18,19^ Congenital anomalies noted in our study area were gastroschisis, omphalocele, trachea-esophageal fistula, encephalocele, VACTERL, spina bifida, Hirschsprung disease, jejunal atresia, pyloric stenosis, anorectal malformation, and imperforate anus. The congenital anomalies consistent with that reported in a tertiary health facility in Niger Delta region by Abolodje et al 2021^17^ are omphalocele, gastroschisis, spinal dysraphism, encephalocele and multiple congenital anomalies (VACTERL).

The distribution of neonatal diseases in our study area differs considerably with that reported in other study areas Neonatal Sepsis – 9.6% (NAUTH) vs 37.6% (ABUTH)^20^, 38.95% (UNIBEN)^21^; Neonatal Jaundice – 5.2% (NAUTH) vs 15% (EKSUTH)^22^ and 35.94% (OKSUTH)^23^; Respiratory Distress Syndrome – 2.6% (NAUTH) vs 58.4% (WGH Ilesa)^24^; Perinatal Asphyxia – 45.0% (NAUTH) vs 11.1% (MASUTH)^25^ and 29.4% (UPTH)^26^. This shows that other centers have higher burden of admissions for neonates with sepsis, jaundice, and respiratory distress syndrome except perinatal asphyxia. The reasons for these disparities could be alluded to the difference in hospital settings and availability of health records. In Fig 1.1, the absence of admissions during the months of August and September 2021 was a consequence of a strike action carried out by the national association of resident doctors, which spanned from August 2nd to October 5^th^, 2021.

**Fig. 1:**
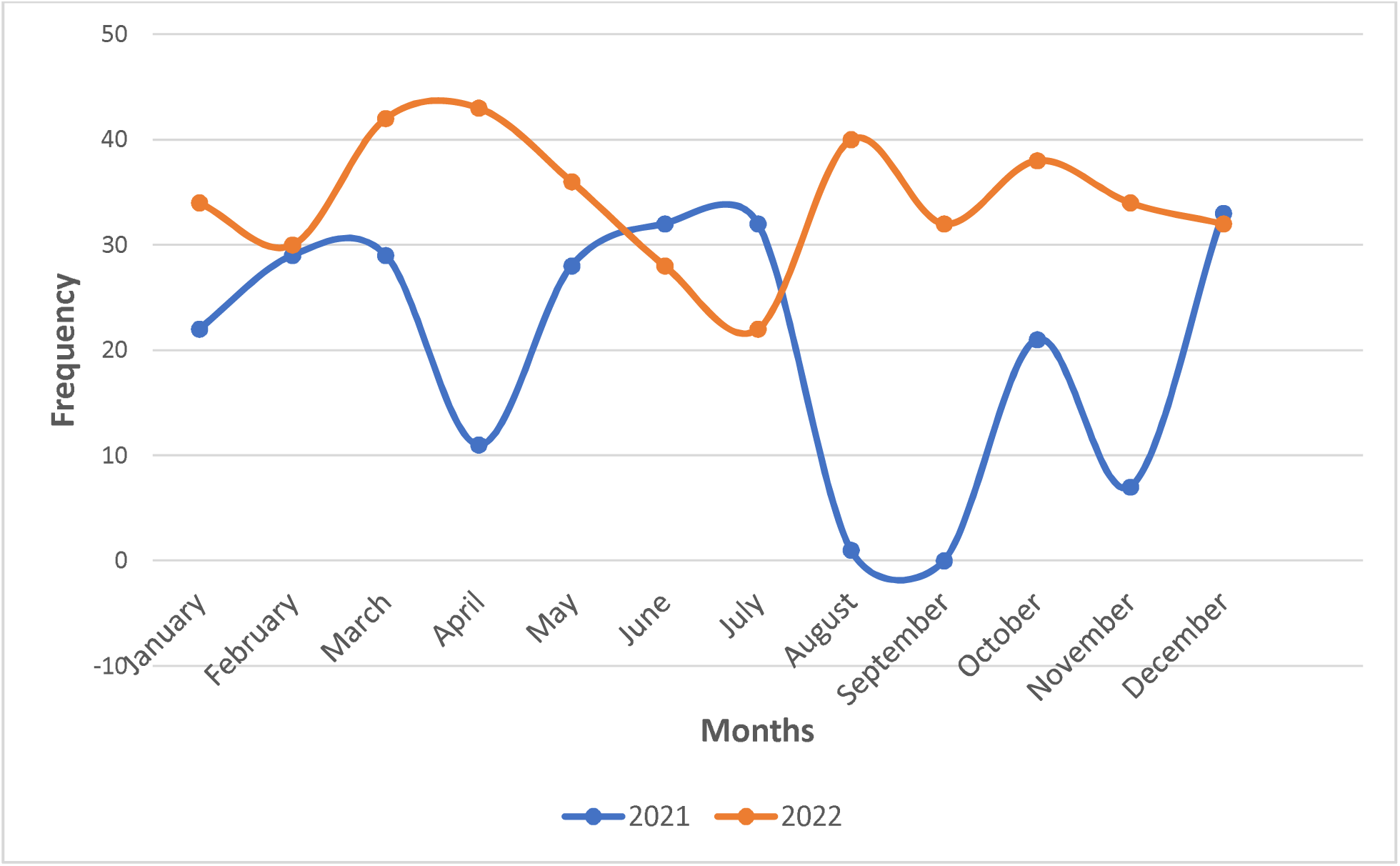
Monthly pattern of neonatal admissions for 2021 and 2022. The comparison of cases between year 2021 and 2022 is shown in supplemental fig. 1. In the year 2021, a higher percentage of cases were seen in the months of June, July, and December. Conversely, the year 2022 had a higher percentage of cases in the other months.

In this study, a higher proportion of admitted neonates (62.8%) were found to be outborn. This trend can be linked to the prevailing preference for traditional birth attendants, local maternities, and non-specialist hospitals, which are perceived as offering more cost-effective childbirth services.^27^ Consequently, neonates are referred to NAUTH when complications arise during childbirth. This finding however differs from findings in other studies conducted at UBTH and ESUTH, where the majority of admitted neonates were inborn.^16,28^ The Implications of this finding are of considerable importance, as it sheds light on the regional disparities in neonatal care and underscores the influence of cultural beliefs on childbirth practices.

Findings from this study show a significant difference in mortality ratio (2:1) between outborn (n=116/412) and inborn (n=33/238). This discrepancy is likely due to the advantages of birth deliveries by skilled health professional, closer scrutiny provided to inborn babies by specialist and experienced doctors, enabling early detection of subtle signs of diseases in newborns. Moreover, the convenience of transferring sick babies delivered in the center to specialized care units (SCBUs) may contribute to the lower mortality rate compared to those born outside the center. The finding is consistent with similar studies conducted in other Nigerian centers, indicating a trend that spans across different locations.^18,28,29^ Most neonatal deaths occurred within the first 24 hours in this study. The finding is consistent with similar studies conducted in other Nigerian centers.^16,30^ This emphasizes the critical need for immediate attention and resuscitation measures to prevent mortality.

As a tertiary institution, 69.2% discharge rate of admitted neonates in our study area poses a concern as to the adequacy of neonatal care efforts. This indicates the need for urgent improvement in the overall neonatal care outcome. Also, the rate of discharge against medical advice (DAMA) was observed to be 4.9% and was attributed to financial constraints and prolonged hospital stay. This finding demonstrates a notable difference when compared to a previous study conducted at UPTH, which reported a DAMA rate of 15%.^29^ The higher DAMA rate observed in UPTH was caregivers’ preference of alternative medicine over continuing hospital treatment due to financial constraints.

Statistically, gestational age at birth, mode of delivery, place of delivery, and duration of hospital stay have shown significant association with neonatal outcome in NAUTH. Our study showed mortality being more likely among premature neonates: <28 weeks as well as outborn neonates. Discharge was more likely among inborn neonates, those delivered by elective caesarean section, those with gestational age of 33-36 weeks and hospital duration between 7-30 days. Neonates with jaundice as well as those delivered by assisted vaginal delivery were more likely to DAMA. Consistent with findings by Nabwera et al^31^, inborn neonates were less likely to die compared to outborn neonates.

## 5. Study Limitations

Data availability in the health records was constrained for the 973 admitted neonates. With just 661 records extracted. Limited data for comprehensive analysis may have introduced outcome bias. Secondary outcome variables (such as caregiver satisfaction and cost of care) could not be evaluated due to the nature of this study.

## 6. CONCLUSION

There is a wide distribution of neonatal diseases in our study area. This phenomenon underscores the critical issue of healthcare accessibility and affordability for specific segments of the population. The study identified associations between care outcomes and factors such as gestational age, delivery mode, birth weight, place of delivery, age at presentation, hospital stay duration. Since mortality was more common in outborn neonates, referral pathways and resources to provide appropriate care need to be prioritized. Despite the comparatively low DAMA rate in our study, it is crucial to recognize that there remains a pressing need to address healthcare disparities and ensure equitable access to medical services. Our study limitations highlight the need to securely keep and store neonatal data for research and clinical purposes. To reduce the burden of neonatal diseases and improve the outcome of neonatal admissions - strengthening perinatal care, emergency obstetric services, and enhancement of neonatal resuscitation skills to traditional birth attendants (TBAs) and other community health workers are key.

## CONTRIBUTORSHIP STATEMENT

Conceptualization: Udochukwu Godswill Anosike, Ugochukwu Godson Amalahu.

Planning: Udochukwu Godswill Anosike, Ugochukwu Godson Amalahu, Chijioke Amara Ezenyeaku, Chika Florence Ubajaka, Anokwulu Ifeanyi Osmond.

Data collation: Udochukwu Godswill Anosike, Ugochukwu Godson Amalahu, Chiamaka Sandra Nsude, Joseph Moses Adeniyi, Chinemerem Okonkwo, Uzoma Love Nwajinka, Chidozie Valentine Akwiwu Uzoma, Chukwuemelie Darlington Okeke.

Data analysis: Anokwulu Ifeanyi Osmond, Malachy Echezona DivineFavour, Udochukwu Godswill Anosike, Ugochukwu Godson Amalahu.

Reporting and Editing: Udochukwu Godswill Anosike, Ugochukwu Godson Amalahu, Chijioke Amara Ezenyeaku, Chika Florence Ubajaka, Malachy Echezona DivineFavour.

## Data Availability

All data produced in the present work are contained in the manuscript.

